# Multidimensional Assurance of an Electronic Health Record Embedded Generative Artificial Intelligence Summarization Tool

**DOI:** 10.64898/2026.08.31.26361660

**Authors:** Pavan Bodanki, Christopher Capone, Nimay S Hazare, Shamay Agaron, Mahima Vijayaraghavan, Sohan Japa, Irina Zaretsky, Jeffrey Epstein, Ashwin Sawant, Ahmed Shaikh, Evan Leibner, Aniket Sharma, Sandeep Gangadharan, Timsal Ghani, Hua-Hsin Tai, Daniel Ngai, Sean El-Haj, Prathamesh Parchure, Annelia Itwaru, Ben Kaplan, Aditi Vakil, Ronald Tamler, Avniel Klein, Robert Freeman, Patricia Kovatch, Bruce Darrow, Jolion McGreevy, Lisa Stump, Girish N Nadkarni, Prem Timsina, Ankit Sakhuja

## Abstract

**Objective:** To evaluate Epic IP Insights, an electronic health record integrated generative AI summarizer used across a seven-hospital health system, using a multidimensional assurance framework.

**Materials and Methods:** We developed and validated an agentic hallucination detector that decomposed summaries into atomic content units (ACUs) and verified each against source documentation. We then applied it to remaining summaries to estimate the hallucination rate. We also measured source note utilization, textual and semantic similarity of regenerated summaries, and clinician perceptions through structured evaluations.

**Results:** The EPIC IP Insights generated 706 summaries across 445 encounters and used a mean of 5.8% of available notes. The detector was developed on 30 summaries (2,012 ACUs) and validated on 5 held out summaries (295 ACUs). The detector agreed with physician adjudication on 96.95% ACUs in the validation set. Across 671 remaining summaries containing 40,452 ACUs, the hallucination rate was 11.79% (95% CI, 11.47% to 12.10%). Among 385 regenerated pairs of summaries, 32.7% were textually identical and 37.9% were semantically identical. Clinicians found the tool easy to use (97.8%), while responses were more mixed regarding reliance on the output with little verification (45.9%), expected efficiency gains (45.3%), and frequent use (46.1%).

**Discussion:** Clinicians found the tool easy to use but believed that outputs required verification. Systematic evaluation identified the frequency of hallucinations and limited use of available notes which provide directions for future improvement.

**Conclusion:** Multidimensional assurance frameworks are needed to evaluate the safety, reliability, and consistency of generative AI tools.

## BACKGROUND AND SIGNIFICANCE

A physician taking over the care for a patient has limited time to review dozens of notes. Chart review is one of the most time-consuming parts of inpatient care, and a major driver of documentation burden. Thus, clinical text summarization has become one of the first serious uses of generative artificial intelligence (AI) in medicine[1]. The early evidence is encouraging. Adapted large language models (LLMs) can summarize clinical text well. Physicians rate their output as equivalent to, or better than, summaries written by medical experts[2]. Electronic health record (EHR) vendors have moved quickly on these results. Generative AI summarization is now built into the chart itself, and health systems are deploying it at scale.

The science of evaluating these tools has advanced quickly alongside them. Prior work has produced frameworks for quantifying hallucinations and clinical safety in LLM-generated medical text[3], instruments for scoring summary quality[4], and methods for tracing each statement back to its source note[1]. Almost all the studies evaluated the models that investigators built or configured themselves, offline, on data they chose. A tool running live in the chart is a different paradigm. It selects its own inputs, and the clinician cannot see what it left out. It can be regenerated on demand, so two clinicians caring for the same patient may not read the same summary. Its errors reach the bedside, not an annotator. A recent study looked at a vendor-built tool inside the EHR where the physicians were broadly positive but reported omissions, confusing content and hallucinations[5]. That evaluation was qualitative feedback from 10 physicians on 147 summaries, with hallucinations identified manually by physicians. Such manual review, however, does not scale to the thousands of summaries a health system generates.

In this study, we evaluate Epic IP Insights, a generative AI-based summarization tool deployed in the live EHR across the seven hospitals within the Mount Sinai Health System, using three complementary approaches. First, we characterized the tool itself, quantifying source note utilization and the reproducibility of repeated summaries generated for the same encounter. Second, we developed a hallucination detector, validated against physician review, that can be applied at scale to the full volume of summaries generated by the tool. Third, we asked practicing clinicians to assess the correctness, safety, and workflow value of the summaries alongside the underlying charts. Together, these analyses close the gap between offline model evaluation and the assurance a health system needs for an EHR-embedded generative AI tool used in routine care.

## MATERIALS AND METHODS

### Study Design

We conducted a retrospective observational study to evaluate the performance of Epic IP Insights for generation of summaries for inpatient encounters at the Mount Sinai Health System (MSHS), a seven-hospital health system in New York City. The evaluation comprised of two components: hallucination analysis, in which we assessed the rate of hallucinations in generated summaries using an agentic pipeline, and a clinician evaluation, in which eight clinicians reviewed summaries and accompanying charts and provided structured ratings.

Prior to the start of the study, the eight clinician evaluators completed a one-hour virtual orientation on the use of the tool. Throughout the study period, we held two weekly office hours (one hour each) for questions, conducted weekly check-ins with all evaluators, and encouraged clinicians to reach out at any time. The evaluators represented a broad range of clinical experience, including Internal Medicine, Emergency Medicine, Pediatrics, and Critical Care.

### Epic IP Insights

Epic IP Insights is a GPT-4[6] powered generative AI–based summarization tool embedded within the Epic electronic health record (EHR). It was introduced at the Mount Sinai Health System (MSHS) on December 1, 2025. The tool ingests longitudinal clinical documentation from a patient encounter and generates a chronological, day-by-day summary of the hospital course, with each day prefaced by its corresponding date. Summaries were generated on demand from a manually triggered dropdown menu within the EHR and could be regenerated every 12 hours. Consequently, a single encounter could be associated with multiple summaries generated at different time points by different users.

The tool uses History and Physical, Emergency Department, Progress, and Consult notes from the current admission. If these are unavailable, it may use discharge summaries from the previous five discharges or office visits from the last year. No explicit note-count, character, or token limit was identified in the available documentation.

### Cohort and Data Extraction

Over a four-week study period (December 1 – December 29, 2025), each week, we randomly identified 25 adult inpatients (age ≥18) discharged from MSHS within the preceding two weeks, with a length of stay between three and seven days and provided their charts to the clinician reviewers. Patients were thus included from November 17 to December 14, 2025. No patient was included more than once across the four weeks, yielding 100 unique charts for clinician evaluation. For the hallucination analysis, we additionally included 345 eligible encounters with available summaries discharged between November 21 to November 29, 2025, which together with 100 clinician reviewed charts yielded a cohort of 445 charts.

### Hallucination Detection Agent

To assess whether the generated inpatient summaries were factually consistent with the underlying clinical record, we implemented an automated hallucination-detection framework based on the decompose-and-verify method of ACUEval[7]. Consistent with the original method, we decomposed each generated summary into discrete, independently verifiable statements, atomic content units (ACUs), and evaluated each statement against the source documentation. We adopted three components of ACUEval without modification: decomposition of the summary into ACUs, independent verification of each ACU, and classification of each ACU as supported (not a hallucination) or unsupported (hallucination). We extended the framework in several ways to address the requirements of longitudinal inpatient clinical documentation. First, our preliminary testing showed that standard decomposition approaches frequently altered clinical meaning through semantic simplification. We, therefore, incorporated clinically specific semantic constraints into both ACU extraction and verification, including preservation of negation, temporality, medication specificity, laboratory values, and uncertainty. Second, as clinically relevant evidence is often distributed across multiple notes and time points during hospitalization, rather than verifying ACUs against a single reference document as commonly performed in ACUEval, we verified each ACU against the complete set of clinical notes provided as input to the inpatient summarization system. Third, we required the verification agent to generate an explicit rationale for each ACU-level determination to improve interpretability, facilitate clinician review, and support clinical auditability. Finally, we validated automated hallucination classifications through a structured clinician review. The resulting framework consisted of two sequential components: (1) an ACU generation agent and (2) an ACU verification agent (Figure 1).

**Figure 1.**
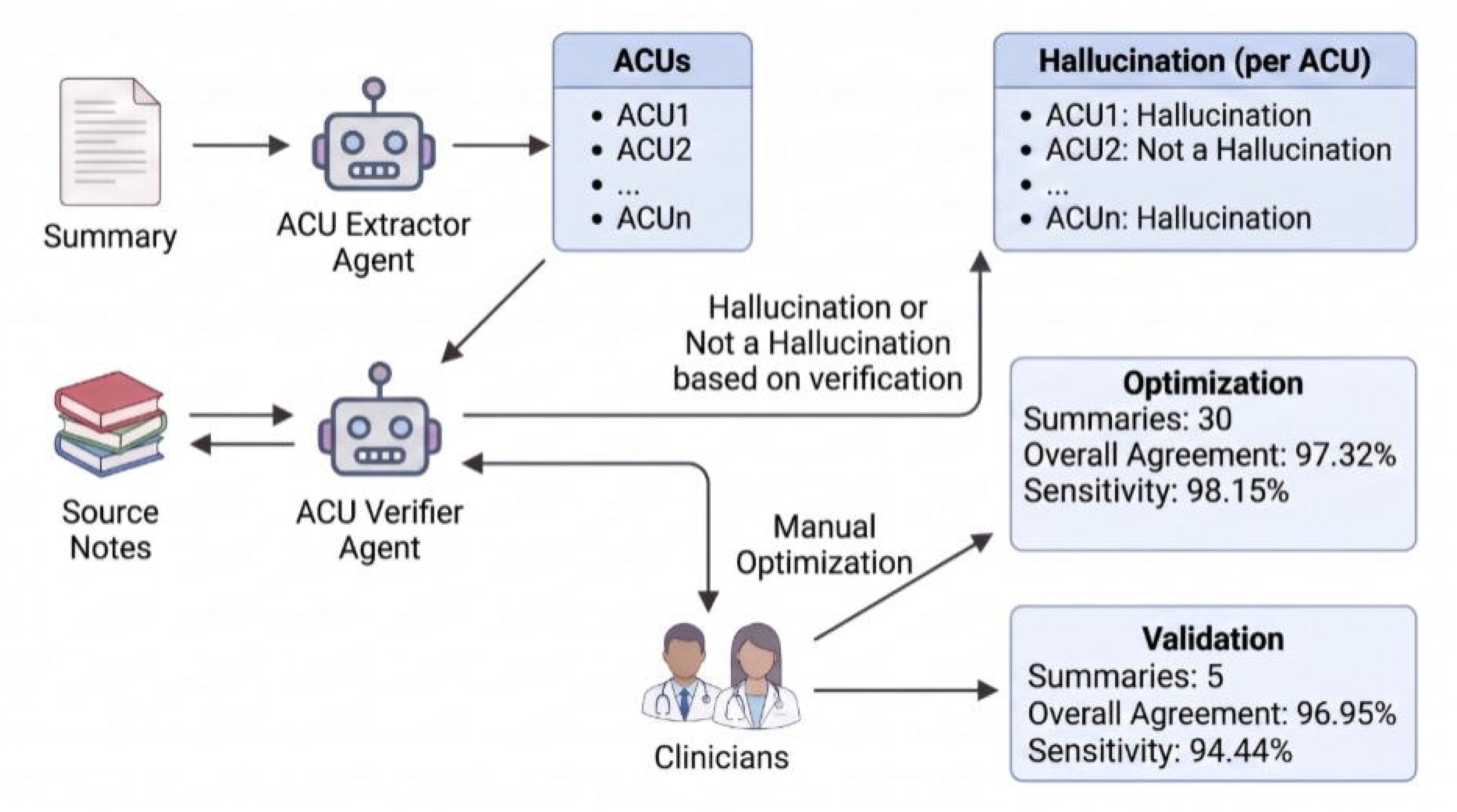
Hallucination Detection Agent. ‘ALT TEXT’ Flowchart illustrates a process for detecting hallucinations in ACUs using an ACU Extractor Agent, ACU Verifier Agent, and clinician input.Key components include data flow from summary and source notes, classification of ACUs as hallucination or not, and performance metrics for optimization and validation phases with overall agreement and sensitivity percentages. This image is generated using biorender.

#### ACU generation agent

We used the ACU generation agent to decompose each generated summary into a set of ACUs. We defined each ACU as a single clinically meaningful and independently verifiable statement. Because inpatient summaries frequently embed several distinct clinical concepts such as diagnoses, procedures, medications, laboratory trends, and temporal events within a single sentence, our objective at this stage was to transform complex narrative summaries into granular semantic units that could be independently verified against source clinical documentation. We generated ACUs using GPT-5.2[8] with an extraction prompt designed for longitudinal inpatient summaries, incorporating explicit rules to preserve clinically important semantic details during decomposition. We instructed the model to follow Table S1.

#### ACU verification agent

We verified each ACU independently using a separate verification agent that classified each ACU as either “Not a hallucination” or a “Hallucination” based on whether sufficient evidence was present within the source documentation. The verification agent used an LLM-based prompt using GPT-5.2[8] designed specifically for clinical factual consistency assessment. The model received the extracted ACU and the complete set of source clinical notes that Epic IP Insights had used to generate the corresponding summary. We designed the verification prompt to evaluate factual consistency across six clinically relevant dimensions (Table 1):

**Table 1.** Clinical Verification Dimensions Considered in ACU Assessment.

| Verification dimension | Explanation |
| --- | --- |
| (1) Negation and polarity. | Checks whether affirmative and negative clinical statements are preserved correctly. A fact should not be reversed (e.g., 'no pneumonia' vs. 'pneumonia'). |
| (2) Agreement of numeric values and associated units. | Ensures all numerical values and measurement units exactly match the source documentation. |
| (3) Medication specificity | Verifies medication identity, dose, route, and frequency whenever these details are available in the source documentation. |
| (4) Temporal alignment between the ACU and source documentation. | Confirms that the timing of events in the ACU matches the timing documented in the source notes. |
| (5) Causal language verification | Accepts causal relationships only when they are explicitly documented rather than inferred. |
| (6) Preservation of diagnostic and procedural specificity. | Ensures diagnoses and procedures retain the same level of specificity as the source documentation. |

**Note:** Each atomic content unit was evaluated independently against the source clinical documentation across six clinically relevant dimensions. ACU, atomic content unit.

### Prompt Optimization and Validation

We performed prompt optimization for both the ACU generation and ACU verification agents iteratively using hallucinated ACUs identified during preliminary testing. For the ACU generation agent, prompt refinement was performed iteratively by analyzing extracted ACUs and introducing constraints to enforce atomicity, preserve clinical qualifiers, and prevent hallucination of unsupported details. Generated ACUs were manually evaluated during preliminary testing to assess whether they were sufficiently granular, faithful to the source summary, clinically complete, and free of unsupported inferences. For example, the summary statement “On 11/21/2025, the patient presented with 3-day persistent headache and intermittent visual flashes, without focal neurological deficits or meningeal signs” was decomposed into four separate ACUs: “On 11/21/2025, the patient presented with 3-day persistent headache,” “On 11/21/2025, the patient had intermittent visual flashes,” “On 11/21/2025, the patient had no focal neurological deficits,” and “On 11/21/2025, the patient had no meningeal signs.” This represents a well-developed ACU set because each statement is clinically meaningful, individually verifiable, temporally anchored, and preserves important qualifiers, including symptom duration and explicitly documented negative findings.

A randomly selected subset of 30 summaries from the study cohort were then used for prompt development and refinement for ACU verification agent. Two reviewers (SA and NH) independently reviewed each ACU from each summary against its source clinical documentation and labeled each clinical statement as hallucination or not a hallucination, establishing the reference standard dataset for hallucinations. Disagreements between the two reviewers were resolved by discussion, with arbitration by a third reviewer (AS) where consensus could not be reached[9]. We then iteratively refined the architecture and prompts of the ACU verification agent to improve its performance till it was able to identify over 95% hallucinations from this reference standard dataset (the sensitivity of the ACU verification agent).

The final verification system comprised three agents: a primary verification agent, a tabular verification agent and a temporal agent. The primary verification agent classified each ACU as hallucinated or not hallucinated against the source notes. A tabular verification agent mapped ACUs to values in tabular evidence stored as flattened text; this agent was necessary because table structure was frequently lost when notes were extracted, leaving rows and columns collapsed into unstructured text that the primary agent could not reliably interpret. A temporal agent flagged ACUs that depended on events not yet occurring at the summary reference time. For example, a summary stating that “initial workup included labs for renal function” was flagged as hallucinated when the notes indicated only a plan (“will obtain labs”) with no subsequent confirmation, anywhere in the input notes, that the labs were performed.

We then evaluated the finalized framework on a held-out validation set of five summaries with 295 ACUs that had not contributed to prompt refinement. Two physicians (TG and HT) reviewed each ACU from each summary against its source notes and labeled the clinical statements as hallucinated or not. Disagreements between the two reviewers were resolved by discussion, with arbitration by a third reviewer (AS) where consensus could not be reached[9]. We then computed agreement between the reviewers’ labels and the framework’s classifications.

### Clinician Evaluation

To assess clinician perceptions of the clinical utility of the tool, each clinician reviewer was asked to rate every assigned summary across ten items on a 5-point Likert scale anchored at Strongly Disagree, Disagree, Neutral, Agree, and Strongly Agree. The questions were as follows in Table S2.

For nine of the ten items, Strongly Agree corresponded to the most favorable response; question 5 was reverse-coded. Reviewers were also asked whether they had personally generated the summary or were reviewing one generated by another user, and were given the option to leave free-text comments.

The weekly sample of 25 visit summaries were randomly and uniformly distributed across the eight clinician evaluators, with each clinician reviewing 10 summaries per week. Over the four-week study period, 100 unique patients were reviewed; 80 were evaluated by three clinicians and 20 by four, yielding a minimum of three independent evaluations per patient. A total of 320 evaluations were completed. Reviewers were blinded to one another’s ratings throughout the study period.

### Statistical Analysis

We summarized continuous variables as mean ± standard deviation (SD) or median [interquartile range(IQR)], depending on distribution, and categorical variables as number and percentage. At the cohort level, we described age-group, sex, race, Charlson Comorbidity Index, hospital site, length of stay, and encounter characteristics. At the summary level, we described the number of summaries generated per encounter, the number of available source notes, the number and type of source notes used by the tool, summary length, and the number of summary statements generated per summary. We evaluated source note utilization rate as the fraction of available documentation that was actually utilized by the tool to generate summaries.

Our primary outcome was the hallucination rate, defined as the proportion of summary statements classified as unsupported by the source documentation. We estimated 95% confidence intervals using a 10,000-iteration cluster bootstrap, resampling. We compared the detector’s classifications against the physician reference standard using overall agreement and sensitivity, with sensitivity capturing whether unsupported clinical statements were correctly identified. We further compared hallucination rates across subgroups defined by age group, sex, and race using Kruskal-Wallis[10] and Mann-Whitney U[11] tests.

To assess reproducibility, we compared regenerated summaries from the same encounter using character-level similarity and BERTScore[12]. Character-level similarity captured textual overlap, whereas BERTScore captured semantic similarity, since summaries may differ in wording while conveying the same clinical meaning. We used Spearman’s correlation[13] to evaluate whether similarity changed as the interval between generations increased.

We summarized clinician survey responses across Likert categories, combining Agree and Strongly Agree as favorable responses. We assessed inter-rater reliability using weighted Gwet’s AC2[14 15], chosen because the ratings were ordinal and expected to concentrate in a few categories, where kappa-based statistics can be unstable, weighting credited partial agreement between adjacent categories.

## RESULTS

### Cohort Characteristics

We analyzed 445 inpatient encounters that met eligibility criteria over the four-week study period. Encounters spanned all seven MSHS hospitals. 53.5% patients were 65 years of age or older, and 58.0% were female. 38.4% of patients were White and 26.1% BlackFull demographic and encounter characteristics are reported in Table 2.

**Table 2.** Demographic Characteristics of the Cohort.

| Variable | Level | Cohort |
| --- | --- | --- |
| **Age group** |  |  |
|  | <65 | 46.5% |
|  | >=65 | 53.5% |
| **Sex** |  |  |
|  | Male | 42.0% |
|  | Female | 58.0% |
| **Race** |  |  |
|  | White | 38.4% |
|  | Black or African American | 26.1% |
|  | Other | 33.0% |
|  | Unknown / missing / declined | 2.5% |
| **Charlson Comorbidity Index** | Mean (SD) | 4.3 (3.6) |

**Note:** Values are presented as percentages unless otherwise indicated. The Charlson Comorbidity Index is presented as mean (standard deviation).

### Characteristics of Summaries

Epic IP Insights generated 706 summaries across the study cohort. For 103 encounters, a summary was generated more than once, producing 385 regenerated summary pairs available for direct comparison; 32.7% of these pairs were exact character-level matches, whereas 67.3% exhibited measurable textual differences.

### Source Note Utilization Rate for Summary Generation

The tool utilized a mean (standard deviation) of 5.8% ± 1% of the notes available at admission per encounter. Note utilization ranged from 0.6% to 39% across encounters Among note types, history and physical notes were utilized most often, in 89.9% of records, followed by ED provider notes (30.9%) and progress notes (30.0%) (Figure 2).

**Figure 2.**
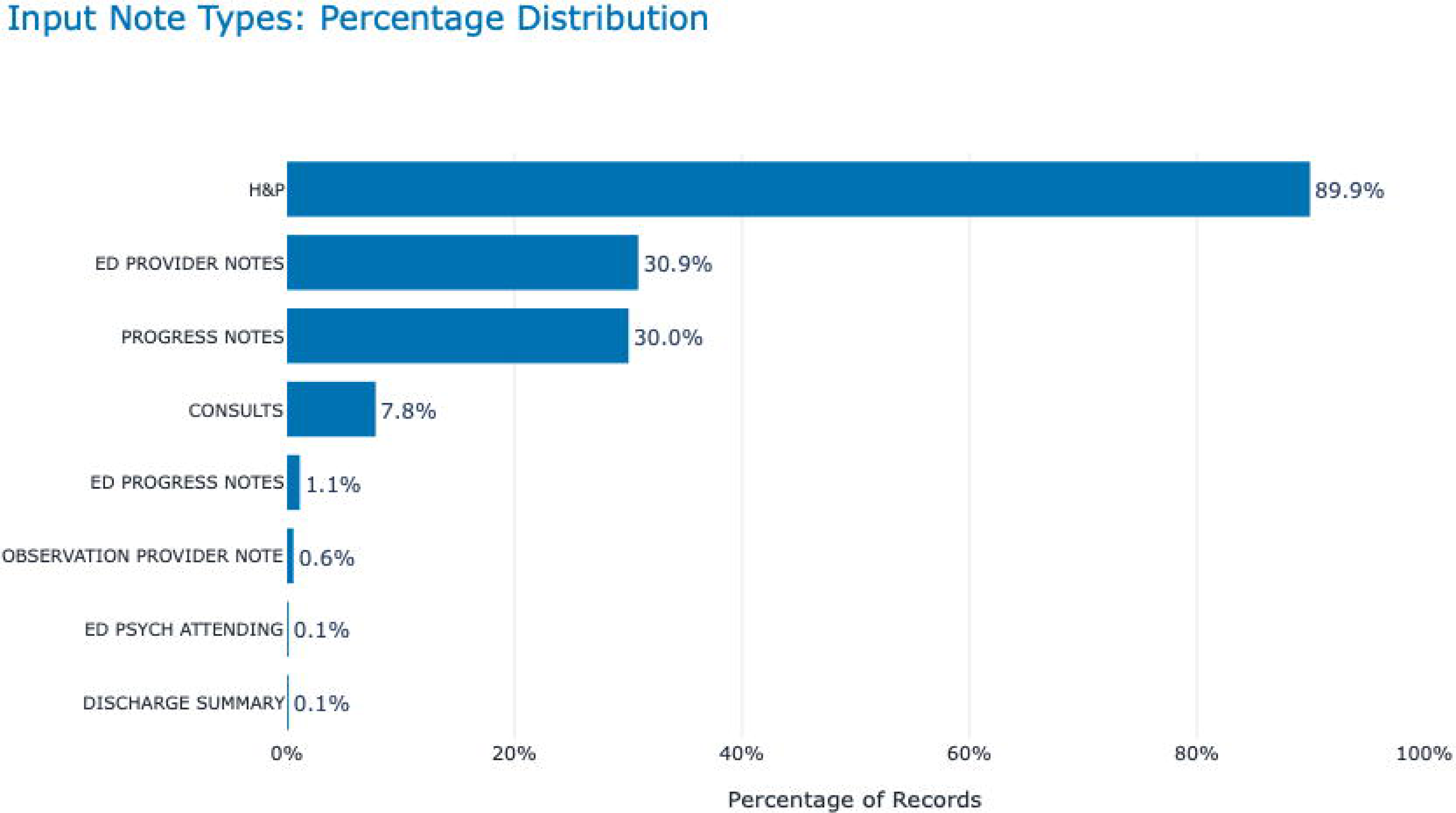
Input Note Type distribution. Bar chart showing percentage distribution of input note types with horizontal blue bars and labels. ‘ALT TEXT’ H&P notes dominate at 89.9%, followed by ED provider notes at 30.9% and progress notes at 30.0%, while other categories remain below 8%.

### Hallucination Rate

On the 30-summary development set, the detector agreed with the physician reviewers (SA and NH) on 97.32% of summary statements and caught 98.15% of the hallucinations they identified. In the held-out test set, physicians identified 18 hallucinations, of which the detector correctly flagged 17 (sensitivity 94.44%). Across all statements in the test set, the detector’s classifications matched the physician reference standard 96.95% of the time, indicating high overall agreement between the detector and clinician judgment.

Having validated the detector against physician judgment, we applied it to the remaining summaries in the cohort. After setting aside those used for model development and validation, 671 summaries remained. The detector extracted 40,452 summary statements from these summaries, of which 4,770 were flagged as hallucinated, a hallucination rate of 11.79% (95% CI 11.47%-12.10%). At the summary level, 654 of 671 summaries (97.5%) contained at least one hallucination. These discrepancies spanned from the mischaracterization of laboratory values, such as confusing hemoglobin with hematocrit, misrepresenting ordered imaging protocols, and falsely asserting that antibiotics were withheld (Table S2).

### Fairness Analysis

No statistically significant differences in hallucination scores were observed across subgroups defined by age group, sex, or race, indicating no evidence of demographic bias in the summarization tool Table S3.

### Reproducibility of Regenerated Summaries

Among non-identical regenerated summary pairs, character-level match declined as the interval between generations lengthened (Spearman’s ρ = −0.456, p < 0.001). On semantic comparison, 37.9% of regenerated pairs were semantically identical (BERTScore = 1.0), and the remainder showed measurable semantic differences. Among pairs with less than 100% character-level match, character overlap was below 50% whenever BERTScore was below 1.0 (Figure 3).

**Figure 3.**
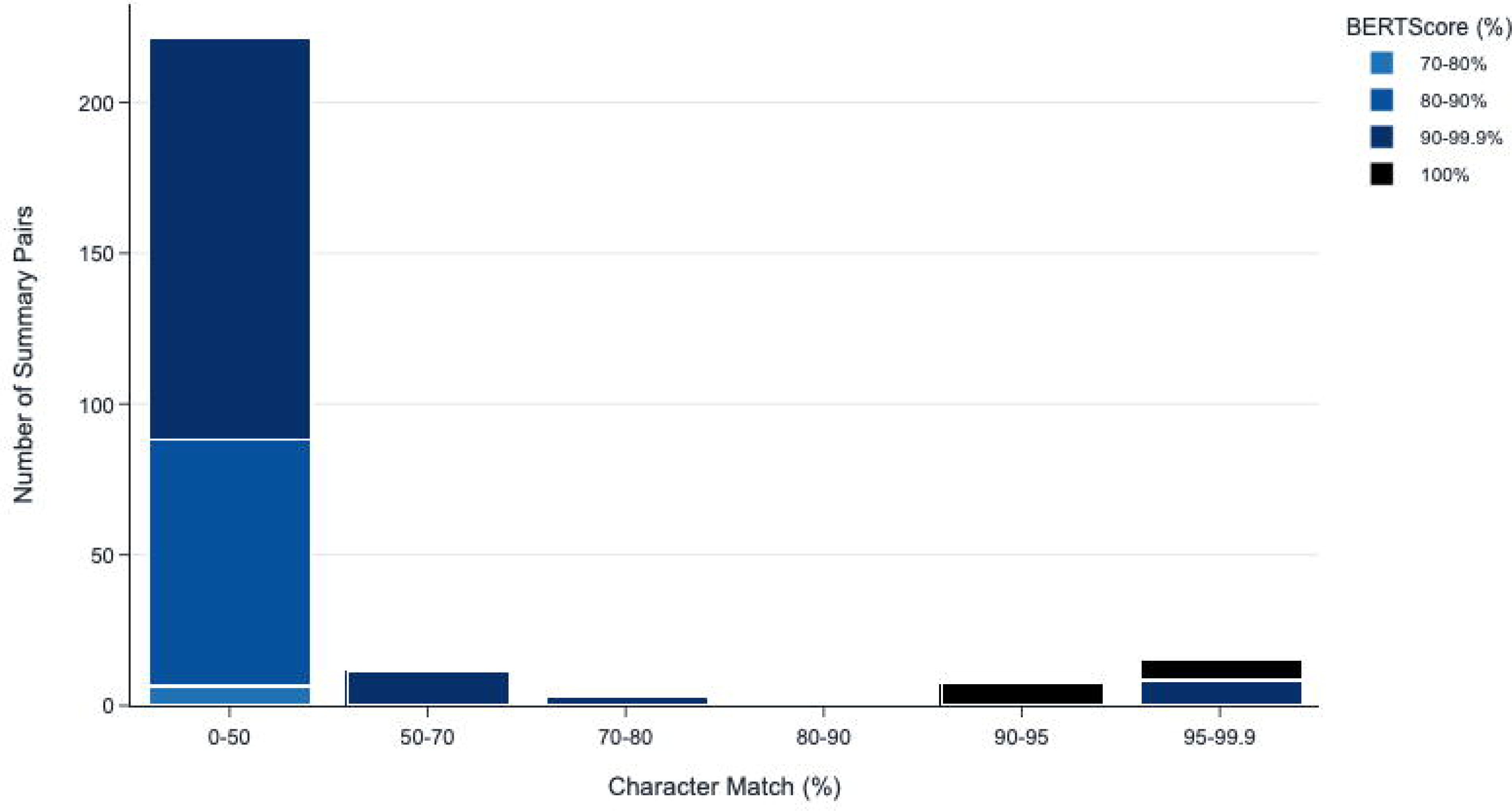
Comparison Between Character Match % and BERT Score. Excluding summaries with 100% character match. ‘ALT TEXT’ Bar chart showing distribution of summary pairs by character match percentage on x-axis and number of summary pairs on y-axis. Color-coded bars represent BERTScore ranges, with most pairs in 0-50% character match and 70-80% BERTScore, and smaller peaks at 90- 95% and 95-99.9% character match with higher BERTScores

### Clinician Evaluation

Clinicians completed 320 evaluations (Table 3) across the 100-chart review subset. Questions related to ease of use (Q9) and absence of hallucinations (Q4) received the most favorable distributions, with 97.8% and 91.3% of responses falling in the Agree or Strongly Agree categories, respectively. Correctness of generated information (Q2) also skewed positively, with 83.1% agreement. Items addressing reliance and workflow integration were rated more conservatively with 45.9% favorable response for whether clinicians could rely on outputs with little additional verification (Q6), 45.3% for whether the tool would improve efficiency (Q8), and 46.1% for whether they would like to use it frequently (Q1, 46.1%). Inter-rater reliability across reviewers was substantial, with a Gwet’s AC2[14 15] of 0.67. Table S4 contains further information on inter-rater reliability.

**Table 3.** Clinician Ratings of the Clinical Utility, Safety, and Usability of the Generative AI Summarization Tool.

| Question | Strongly Disagree | Disagree | Neutral | Agree | Strongly Agree |
| --- | --- | --- | --- | --- | --- |
| Q1: I would like to use this tool frequently | 2.82%<br>(9) | 19.12%<br>(61) | 31.97%<br>(102) | 34.48%<br>(110) | 11.60%<br>(37) |
| Q2: The AI tool generated information was correct | 2.19%<br>(7) | 10.63%<br>(34) | 4.06%<br>(13) | 69.06%<br>(221) | 14.06%<br>(45) |
| Q3: The AI tool generated information reliably captured all key details | 6.25%<br>(20) | 19.69%<br>(63) | 5.63%<br>(18) | 41.25%<br>(132) | 27.19%<br>(87) |
| Q4: The AI tool outputs were free from false or fabricated information (hallucinations) | 1.25%<br>(4) | 5.31%<br>(17) | 2.19%<br>(7) | 62.19%<br>(199) | 29.06%<br>(93) |
| Q5: The AI tool's output could pose a risk to patients or organizational processes if used without verification | 11.88%<br>(38) | 39.06%<br>(125) | 16.56%<br>(53) | 30.63%<br>(98) | 1.88%<br>(6) |
| Q6: I could rely on the GenAI tool's outputs with little need for additional verification | 5.31%<br>(17) | 37.50%<br>(120) | 11.25%<br>(36) | 41.88%<br>(134) | 4.06%<br>(13) |
| Q7: I felt confident using this AI tool in my workflow | 2.50%<br>(8) | 20.63%<br>(66) | 15.63%<br>(50) | 53.13%<br>(170) | 8.13%<br>(26) |
| Q8: I am confident that this AI tool will improve my efficiency | 3.44%<br>(11) | 35.94%<br>(115) | 15.31%<br>(49) | 37.19%<br>(119) | 8.13%<br>(26) |
| Q9: It was easy to learn how to use this AI tool effectively | 0%<br>(0) | 1.56%<br>(5) | 0.63%<br>(2) | 59.69%<br>(191) | 38.13%<br>(122) |
| Q10: The AI tool's outputs were presented clearly | 2.19%<br>(7) | 9.38%<br>(30) | 10.94%<br>(35) | 46.56%<br>(149) | 30.94%<br>(99) |

**Note:** Values are presented as number of clinician evaluations in each response category. Ratings were collected using a 5-point Likert scale. A total of 320 clinician evaluations were completed across 100 unique patient charts. Counts for Q1 sum to 319 because one response was missing. For all items except Q5, higher ratings indicate more favorable perceptions of the tool. Q5 is negatively worded and should be reverse-coded when calculating a composite or favorable-response score. GenAI, generative artificial intelligence.

## DISCUSSION

This evaluation of a live, EHR-embedded generative AI summarization tool across seven hospitals demonstrates the value of examining multiple dimensions of performance after clinical implementation. Clinician evaluations were generally positive for correctness and ease of use, but more cautious about relying on the output without verification, expecting it to improve efficiency, and using it frequently. Our structured analysis similarly identified areas in which factual consistency, source note utilization, and reproducibility could be improved. Together, these findings suggest that clinician perceptions and structured assessments provide complementary dimensions of assurance.

The difference between the 91.3% of clinician ratings indicating that summaries were free from fabricated information and the 11.79% hallucination rate identified through statement by statement review may reflect differences in the mode of review. In routine clinical workflows, clinicians are likely to assess whether a summary is coherent, plausible, and useful for orientation rather than independently verify every statement against the underlying record. Their simultaneous reluctance to rely on the summaries with little additional verification suggests that clinicians recognized limitations in the outputs even when they could not consistently identify or localize individual discrepancies. This pattern supports the inclusion of both end user feedback and structured factual consistency assessment in postimplementation assurance. Similar differences have been observed in prior studies. Clinicians using an EHR integrated chart review tool spontaneously reported five hallucinations across 147 summaries[5], whereas a structured review of GPT-4 generated emergency department summaries identified hallucinations in 42% of outputs[16]. Although these studies evaluated different tools, settings, and summary types, together they suggest that observed hallucination rates may depend partly on the granularity and structure of the review process.

Our validation findings provide direct support for this interpretation. When physicians performed focused review of individual atomic content units against the source documentation, their classifications closely aligned with those of the agentic hallucination detector. This suggests that the apparent difference between routine clinician ratings and structured audit findings was not primarily due to disagreement about what constitutes an error, but rather to differences in the mode and granularity of review. Automated methods may therefore complement clinician evaluation by enabling more granular and scalable assessment. Beyond factual consistency, source note utilization represents another important dimension of assurance. Under the evaluated configuration, the tool used a small proportion of available notes and relied most often on History and Physical documentation, which may affect completeness as the clinical course evolves. Prior studies have identified omissions in AI summarization tools, including 46 omissions across 147 summaries in an EHR integrated summarization tool[5], and clinically relevant omissions in 47% of GPT 4 generated emergency department summaries[16]. Although note utilization is not a direct measure of omission, these findings support improving source selection, provenance, and transparency regarding which notes contributed to each summary.

Reproducibility emerged as another important dimension of assurance. Among 385 regenerated summary pairs from the same encounter, only 32.7% were textually identical and 37.9% were semantically identical. Textual similarity also declined as the interval between generations increased (Spearman’s ρ = −0.456, p < 0.001), indicating that repeated generation from the same encounter frequently produced materially different outputs. Although our analysis did not determine whether these differences altered the represented clinical course, the extent of variability is important because clinicians may encounter different summaries of the same hospitalization. Future studies should directly assess whether such variation is primarily stylistic or affects clinically relevant content and interpretation.

Clinician feedback showed measured enthusiasm with a clear note of caution. 97.8% of evaluations agreed the tool was easy to use and 83.1% judged the generated information as correct, but clinicians were more reserved about relying on the output without verification, expecting efficiency gains, or using it frequently. Specifically, 45.9% of responses supported reliance with little additional verification, 45.3% supported improved efficiency, and 46.1% supported frequent use. These patterns suggest that clinicians saw value in the tool while also identifying limits that can guide refinement, implementation support, and additional safeguards. More broadly, vendors should disclose the tool’s observed error rate directly within the interface, such as the hallucination rate or another clinically meaningful measure of factual error, so clinicians can see it at the point of use and calibrate their trust accordingly.

Our study has several limitations that must be acknowledged. First, we classified statements as hallucinations or not without grading clinical severity, so our rate mixes consequential errors with trivial ones, and we cannot say how many would have altered management. Second, we did not measure omissions directly, though the low note utilization we observed suggests they are common. Third, our detector verified summaries against the notes the tool ingested rather than the full record. We regard this as the appropriate standard, since a tool cannot assert what it was not given, but it means a statement identified as a hallucination may occasionally reflect a fact documented elsewhere in the chart. Fourth, our cohort was limited to adult inpatients with a length of stay of three to seven days, and performance in pediatric patients or in longer, more complex hospitalizations, where note utilization is likely to be lower still, remains uncharacterized. Finally, we evaluated one vendor-built tool at one health system over four weeks, which limits generalizability to other configurations, prompting pipelines, and subsequent model versions.

## CONCLUSION

These findings support the need for automated, scalable, multidimensional assurance frameworks to monitor safety, reliability, and reproducibility in real world clinical use. Such frameworks, together with end user feedback, should inform iterative refinements in source selection, provenance, and workflow integration. They also highlight the importance of close collaboration between vendors and health systems as generative AI tools become increasingly commonplace in the EHR.

## DATA SHARING STATEMENT

The data underlying this study consist of protected health information contained within electronic health records of patients treated at the Mount Sinai Health System and cannot be publicly shared because of patient privacy, institutional, and regulatory restrictions. De- identified aggregate data supporting the findings of this study may be made available from the corresponding author upon reasonable request and subject to approval by the Mount Sinai Health System and applicable data use agreements.

## Supporting information

Supplementary Material: Additional Explanations, Tables, and Figures

Tripod LLM checklist

## Data Availability

The data underlying this study consist of protected health information contained within electronic health records of patients treated at the Mount Sinai Health System and cannot be publicly shared because of patient privacy, institutional, and regulatory restrictions. De-identified aggregate data supporting the findings of this study may be made available from the corresponding author upon reasonable request and subject to approval by the Mount Sinai Health System and applicable data use agreements.

## REFERENCES

1. Verma R, Alsentzer E, Strasser Z, et al. Verifiable Summarization of Electronic Health Records Using Large Language Models to Support Chart Review. medRxiv 2025 doi: 10.1101/2025.06.02.25328807 [published Online First: 20250603].

2. Van Veen D, Van Uden C, Blankemeier L, et al. Adapted large language models can outperform medical experts in clinical text summarization. Nat Med 2024;30(4):1134–42 doi: 10.1038/s41591-024-02855-5 [published Online First: 20240227].

3. Asgari E, Montaña-Brown N, Dubois M, et al. A framework to assess clinical safety and hallucination rates of LLMs for medical text summarisation. NPJ Digit Med 2025;8(1):274 doi: 10.1038/s41746-025-01670-7 [published Online First: 20250513].

4. Croxford E, Gao Y, Pellegrino N, et al. Development and validation of the provider documentation summarization quality instrument for large language models. J Am Med Inform Assoc 2025;32(6):1050–60 doi: 10.1093/jamia/ocaf068.

5. Kahl N, Frieden M, Pope Z, et al. Evaluation of electronic health record-integrated artificial intelligence chart review. npj Health Systems 2026;3 doi: 10.1038/s44401-025-00064-x.

6. OpenAI. OpenAI. Introducing GPT-4. Secondary OpenAI. Introducing GPT-4 2023. https://openai.com/index/gpt-4-research/?hl=en-IN.

7. ACUEval: Fine-grained Hallucination Evaluation and Correction for Abstractive Summarization; 2024 August; Bangkok, Thailand. Association for Computational Linguistics.

8. OpenAI. OpenAI. Introducing GPT-5-2. Secondary OpenAI. Introducing GPT-5-2 2026. https://openai.com/index/introducing-gpt-5-2/.

9. Akl E, Altman D, Aluko P, et al. Cochrane Handbook for Systematic Reviews of Interventions, 2019.

10. Kruskal WH, Wallis WA. Use of Ranks in One-Criterion Variance Analysis. Journal of the American Statistical Association 1952;47(260):583–621 doi: 10.1080/01621459.1952.10483441.

11. Mann HB, Whitney DR. On a Test of Whether one of Two Random Variables is Stochastically Larger than the Other. The Annals of Mathematical Statistics 1947;18(1):50–60 doi: 10.1214/aoms/1177730491.

12. Zhang T, Kishore V, Wu F, Weinberger KQ, Artzi Y. BERTScore: Evaluating Text Generation with BERT. 2019. https://ui.adsabs.harvard.edu/abs/2019arXiv190409675Z (accessed April 01, 2019).

13. Spearman C. The proof and measurement of association between two things. By C. Spearman, 1904. Am J Psychol 1987;100(3-4):441–71.

14. Landis JR, Koch GG. The measurement of observer agreement for categorical data. Biometrics 1977;33(1):159–74.

15. Gwet K. Handbook of inter-rater reliability: The definitive guide to measuring the extent of agreement among raters, 2012.

16. Williams CYK, Bains J, Tang T, et al. Evaluating large language models for drafting emergency department encounter summaries. PLOS Digital Health 2025;4(6):e0000899 doi: 10.1371/journal.pdig.0000899.

