## Supplementary Material: Additional Explanations, Tables, and Figures for "Multidimensional Assurance of an Electronic Health Record Embedded Generative Artificial Intelligence Summarization Tool"

**Supplementary Material for Multidimensional Assurance of an Electronic Health Record Embedded Generative AI Summarization Tool**

**Supplementary Tables:**

Table S1. Atomic Clinical Unit (ACU) Generation Criteria

(1) Generate one clinically verifiable statement per ACU.

(2) Split compound sentences into separate ACUs when multiple independent claims were present.

(3) Preserve negation and uncertainty terms (eg, “denies,” “no evidence of,” “possible,” “concern for”).

(4) Preserve temporal qualifiers including dates, admission context, procedural timing, and repeated events.

(5) Preserve numeric values, units, and medication-specific attributes including dose, route, and frequency when explicitly present in the summary.

(6) avoid introducing information not explicitly stated in the generated summary.

**Note:** Each generated summary was decomposed into independently verifiable ACUs. Negation, uncertainty, temporal context, numeric details, and medication attributes were preserved exactly as stated. No information absent from the generated summary was introduced.

Table S2. Representative Hallucinated Atomic Content Units and Clinician Review Rationales

| **Claim in Generated Summary** | **Reason provided by Clinicians** |
| --- | --- |
| **Lab test** | |
| The patient summary mentioned Hemoglobin was 37.1 | Hemoglobin level was not ordered for the patient; Hematocrit level was 37.1; the LLM mistook Hematocrit for Hemoglobin |
| The patient summary mentioned Troponin test results are pending | Troponin levels were documented to be negative in History and Physical notes |
| The patient summary mentioned that Complete Blood Count was unremarkable and all parameters were within normal limits | The Complete Blood Count reports documented a low platelet count, and high Mean Corpuscular Volume, Mean Platelet Volume and monocyte counts |
| The patient summary mentioned that Blood culture reports were pending | The notes documented that Urine culture reports were pending, not blood culture |
| The patient summary mentioned that Arterial Blood Gas showed pH of 7.38 | The sample drawn was a venous sample with venous pH 7.38 |
| The patient summary mentioned that the basic metabolic panel reports were unremarkable | The notes documented a high sodium, potassium and chloride levels |
| The patient summary mentioned that the basic metabolic panel reports were unremarkable | The notes documented a high Blood Urea Nitrogen and creatinine level |
| **Reports** | |
| The patient summary mentioned that 2 units Red Blood Cells were transfused to the patient | The History & Physical note documented that 2 units Red Blood Cells were ordered but there was documentation supporting transfusion of only 1 unit Red Blood Cells |
| The patient summary mentioned that contrast enhanced CT abdomen/pelvis was ordered to evaluate patient's intra-abdomen infection | The clinical note documented that CT abdomen/pelvis without contrast was ordered to evaluate patients’ kidneys and screen the intra-abdominal infection |
| The patient summary mentioned that the reason for patient's admission was Oxygen requirement | The notes documented that the patient did have a fall in oxygen levels but the reason for admission was an episode of atrial fibrillation with RVR during dialysis due to suspected pulmonary embolism |
| The patient summary mentioned that the reason for patient's admission was Influenza A | The notes documented that the patient was admitted for suspected pulmonary embolism/tachycardia, while Influenza A was an incidental finding after admission |
| The patient summary mentioned that abdominal distension was evaluated with CT Abdomen with Intravenous contrast to rule out shortness of breath | The notes documented that there is a plan to perform CT Abdomen/Pelvis but the radiological test was not performed during this hospital encounter. |
| The patient summary mentioned that during the patient's past hospitalization, the patient was treated with Levaquin. | The notes documented that the patient refused admission on the dates mentioned in patient summary |
| The patient summary mentioned that the reports of CT pelvis with Intravenous contrast showed no air | The notes documented that CT pelvis with Intravenous contrast performed on given dates reported air and fluid collection |
| The patient summary mentioned that the patient's discharge plan includes monitoring of pneumothorax | The notes documented that the discharge plan did not mention monitoring of pneumothorax |
| **Medications** | |
| The patient summary mentioned that a home-medication (methotrexate) was held | The patient notes documented holding of mycophenolate mofetil and metformin but does not mention withholding methotrexate |
| The patient summary mentioned that the patient was transitioned to another antibiotic (ceftriaxone) | The patient notes documented that the antibiotic (ceftriaxone) was not given to the patient |
| The patient summary mentioned that the medication (pantoprazole) was started between the given dates | The notes documented that the medication (pantoprazole) was part of the plan, but it was not started between the given dates |
| The patient summary mentioned that the patient was not given any antibiotic support since there was no evidence of infection | The patient notes documented that an empiric antibiotic was given to the patient |
| The patient summary mentioned potassium chloride (CR 20 mEq) as being taken daily by the patient as a part of his home medication | The patient notes documented that the medication was prescribed to patient in the Hospital and was not a part of home medication |
| The patient summary mentioned that the medication furosemide 40mg was initiated in this admission for management of volume overload (Heart Failure), elevated BNP and obesity | The patient note mentioned BNP was not elevated, and furosemide was a home medication that was continued, not initiated afresh during admission |
| The patient summary mentioned that medication (prednisone) was patient’s active home medication | The patient notes documented that the patient was not taking medication (prednisone) at home |
| **Symptoms** | |
| The patient summary mentioned that the patient has fever | The patient note documented the highest recorded temperature of 37.6°C (99.7°F) |
| The patient summary mentioned that the patient's admission was complicated by persistent headache | The patient notes documented headache as one of the complains by the patient but admission was due to Ulcerative Colitis flare accompanied by rectal bleeding |
| **General** | |
| The patient summary mentioned that the patient's case was discussed with neurology | The note documented that the case was discussed with the Attending physician in the Emergency Department and the patients Primary Care Physician, but does not mention any Neurology Physician discussion |

**Note:** Representative hallucinated claims are grouped by clinical domain. Claims in the generated summary were compared with the corresponding source clinical documentation. The reasons provided by clinicians describe why each claim was considered hallucinated, including factual inaccuracies, unsupported information, or inconsistencies with the source record.

| **Category** | **Group1** | **#Group1** | **Group2** | **# Group2** | **P-Value** | **P_Adj_Bonferroni** | **Significant** |
| --- | --- | --- | --- | --- | --- | --- | --- |
| **Age** | 18-44 | 124 | 65+ | 365 | 0.0073 | 0.0218 | Yes |
|  | 45-65 | 169 | 65+ | 365 | 0.0272 | 0.0816 | No |
|  | 18-44 | 124 | 45-65 | 169 | 0.4374 | 1 | No |
| **Gender** | Female | 365 | Male | 293 | 0.6097 | 0.6097 | No |
| **Ethnicity** | Other | 187 | White | 254 | 0.0119 | 0.0712 | No |
|  | ASIAN | 22 | White | 254 | 0.0979 | 0.5876 | No |
|  | ASIAN | 22 | Black or African-American | 22 | 0.3842 | 1 | No |
|  | ASIAN | 22 | Other | 187 | 0.7398 | 1 | No |
|  | Black or African-American | 22 | Other | 187 | 0.4234 | 1 | No |
|  | Black or African-American | 22 | White | 254 | 0.8408 | 1 | No |

Table S3. Fairness Analysis Across Demographic Groups Using Pairwise Mann-Whitney U Tests

**Note:** Pairwise differences in hallucination rates were assessed using the **Mann-Whitney U test** with **Bonferroni correction** for multiple comparisons. Statistical significance was defined as an adjusted p-value < 0.05. Only the comparison between the 18-44 and 65+ age groups was significant; however, this finding should be interpreted cautiously due to unequal group sizes. No significant differences were observed across gender or ethnicity groups after adjustment.

Table S4. Inter – Rater Reliability clinician ratings by questions

| Question | Score | Meaning |
| --- | --- | --- |
| 1 | 0.6481 | Substantial |
| 2 | 0.8182 | Almost Perfect |
| 3 | 0.4479 | Moderate |
| 4 | 0.8551 | Almost Perfect |
| 5 | 0.5505 | Moderate |
| 6 | 0.5949 | Moderate |
| 7 | 0.7144 | Substantial |
| 8 | 0.6565 | Substantial |
| 9 | 0.9136 | Almost Perfect |
| 10 | 0.6544 | Substantial |

**Note:** Inter-rater reliability was estimated using weighted Gwet’s AC2 to account for the ordinal 5-point Likert response scale. Agreement strength was interpreted using the Landis and Koch benchmarks: slight, 0.00-0.20; fair, 0.21-0.40; moderate, 0.41-0.60; substantial, 0.61-0.80; and almost perfect, 0.81-1.00. AC2, agreement coefficient 2; Q, survey question.
